# Transmission potential of vaccinated and unvaccinated persons infected with the SARS-CoV-2 Delta variant in a federal prison, July—August 2021

**DOI:** 10.1101/2021.11.12.21265796

**Authors:** Phillip P. Salvatore, Christine C. Lee, Sadia Sleweon, David W. McCormick, Lavinia Nicolae, Kristen Knipe, Thomas Dixon, Robert Banta, Isaac Ogle, Cristen Young, Charles Dusseau, Shawn Salmonson, Charles Ogden, Eric Godwin, TeCora Ballom, Tara Ross, Nhien Tran Wynn, Ebenezer David, Theresa K. Bessey, Gimin Kim, Suganthi Suppiah, Azaibi Tamin, Jennifer L. Harcourt, Mili Sheth, Luis Lowe, Hannah Browne, Jacqueline E. Tate, Hannah L. Kirking, Liesl M. Hagan

## Abstract

**Background:** The extent to which vaccinated persons who become infected with SARS-CoV-2 contribute to transmission is unclear. During a SARS-CoV-2 Delta variant outbreak among incarcerated persons with high vaccination rates in a federal prison, we assessed markers of viral shedding in vaccinated and unvaccinated persons.

**Methods:** Consenting incarcerated persons with confirmed SARS-CoV-2 infection provided mid-turbinate nasal specimens daily for 10 consecutive days and reported symptom data via questionnaire. Real-time reverse transcription-polymerase chain reaction (RT-PCR), viral whole genome sequencing, and viral culture was performed on these nasal specimens. Duration of RT-PCR positivity and viral culture positivity was assessed using survival analysis.

**Results:** A total of 978 specimens were provided by 95 participants, of whom 78 (82%) were fully vaccinated and 17 (18%) were not fully vaccinated. No significant differences were detected in duration of RT-PCR positivity among fully vaccinated participants (median: 13 days) versus those not fully vaccinated (median: 13 days; p=0.50), or in duration of culture positivity (medians: 5 days and 5 days; p=0.29). Among fully vaccinated participants, overall duration of culture positivity was shorter among Moderna vaccine recipients versus Pfizer (p=0.048) or Janssen (p=0.003) vaccine recipients.

**Conclusions:** As this field continues to develop, clinicians and public health practitioners should consider vaccinated persons who become infected with SARS-CoV-2 to be no less infectious than unvaccinated persons. These findings are critically important, especially in congregate settings where viral transmission can lead to large outbreaks.

## Introduction

COVID-19 vaccines are highly effective in preventing severe illness and death from SARS-CoV-2 (the virus that causes COVID-19). However, because COVID-19 vaccines are not 100% effective in preventing infection, some infections among vaccinated persons are expected to occur. As global vaccination coverage increases, the role of vaccinated persons in transmission will be a critical determinant of the pandemic’s future trajectory.^1^ The extent to which vaccinated persons who become infected contribute to transmission of SARS-CoV-2, including the B.1.617.2 (Delta) variant, is not yet well understood. Some preprint manuscripts have reported comparable indicators of transmission potential regardless of vaccination status,^2^ while others have reported reduced viability of virus isolated from vaccinated persons.^3^

The Delta variant has been associated with a peak in COVID-19 cases in the United States beginning in July 2021 that included large outbreaks among vaccinated and unvaccinated persons in crowded settings.^4-6^ These findings are of particular concern for congregate living environments such as correctional and detention facilities and long-term care facilities because of the potential for rapid transmission of SARS-CoV-2 and the high prevalence of underlying health conditions associated with severe COVID-19.^7-9^

In a recent outbreak involving the Delta variant in a federal prison in Texas, the cumulative incidence of infection in two affected housing units was 74%; it was 93% and 70% among unvaccinated and vaccinated incarcerated persons, respectively.^6^ Using serial mid-turbinate nasal specimens collected from a subset of incarcerated persons infected during this outbreak, this report assesses reverse transcription-polymerase chain reaction (RT-PCR) and viral culture characteristics as surrogate markers of transmission potential among persons fully vaccinated and those not fully vaccinated over time. This report is one of the first longitudinal investigations of viral shedding from vaccinated persons infected with the Delta variant and contributes to the evidence base guiding infection prevention and control procedures across a variety of settings.

## Methods

### Investigational Setting

On July 12, 2021, an outbreak of SARS-CoV-2 among vaccinated and unvaccinated persons was detected in a federal prison in Texas. Staff from the Centers for Disease Control and Prevention (CDC) and Federal Bureau of Prisons (BOP) deployed to the prison to investigate the outbreak as previously reported.^6^ As part of this outbreak investigation, a subset of incarcerated persons provided serial mid turbinate nasal specimens which were analyzed to evaluate the potential role of infected vaccinated and unvaccinated persons in transmission of SARS-CoV-2. This activity was reviewed and approved by the BOP Research Review Board and CDC and conducted consistent with applicable federal law and CDC policy.^*^

### Participant Enrollment and Serial Specimen Collection

Incarcerated persons living in four housing units where COVID-19 cases had been identified were invited to participate in serial swabbing. Persons were eligible to enroll if they had tested positive for SARS-CoV-2 between July 12 (the start of the outbreak) and August 4, 2021. CDC and BOP staff held information sessions to explain the purpose of the project and to answer questions, including privacy protections and how results of the study would be made available to participants. All persons choosing to participate signed informed consent forms, which were provided in English and Spanish.

Specimen collection occurred during July 18—August 9, 2021. CDC and BOP staff collected one nasal mid-turbinate specimen daily for 10 consecutive days from participants who had tested positive, beginning on July 19 or, for cases identified after July 19, beginning on the date of participants’ first positive test. All incarcerated persons residing in housing units where cases were identified were placed under quarantine precautions. To assist in case-finding, consenting persons who were quarantined were tested every other day beginning on July 19 or on their first full day of quarantine; those who tested positive during quarantine were invited to participate in the 10 consecutive days of specimen collection. All participants were asked to provide a specimen on August 6 to provide data additional data on viral shedding, which corresponds to a late timepoint in infection for most participants (Figure 1).

**Figure 1.**
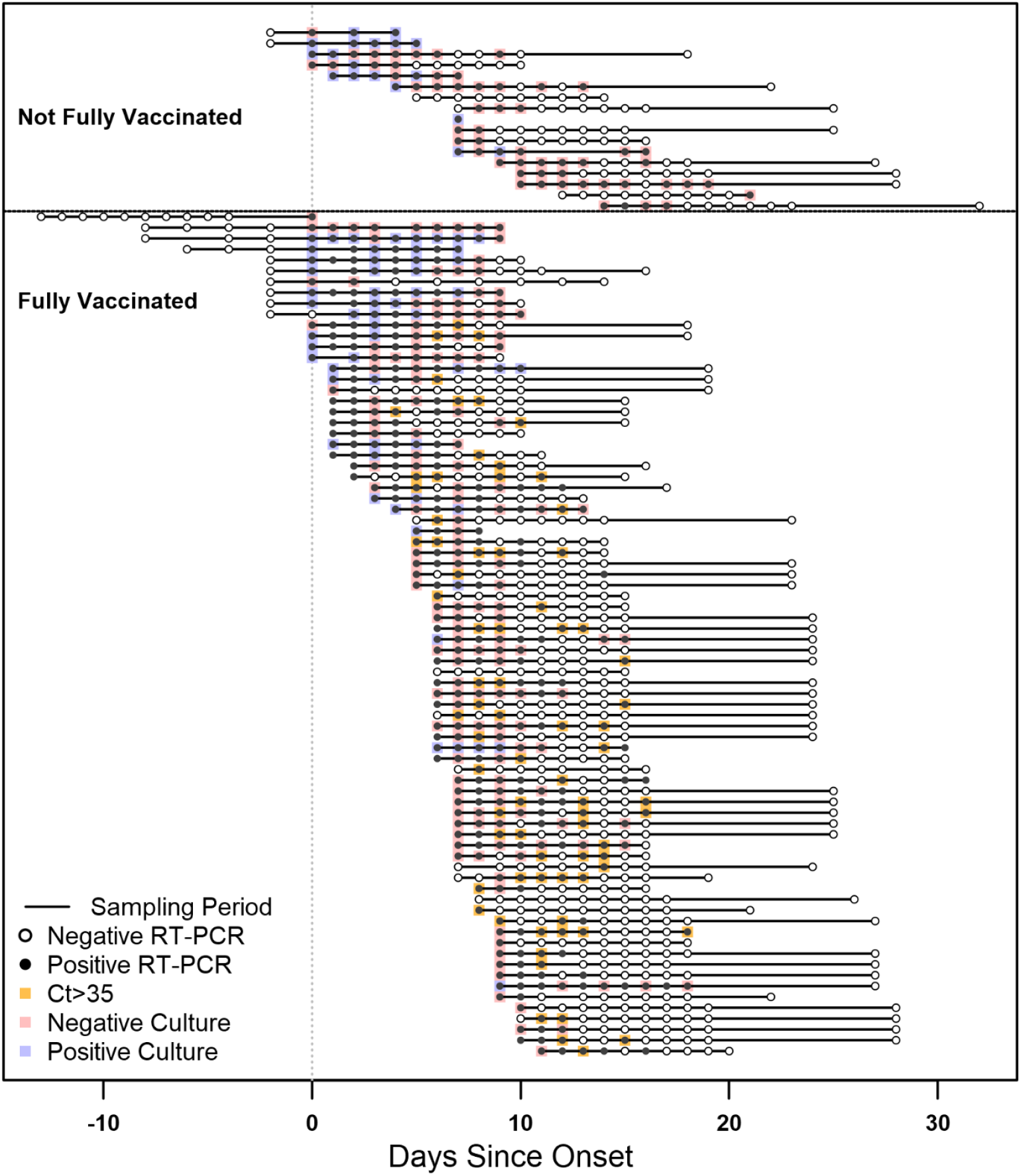
Timelines and results of nasal mid-turbinate specimens collected from enrolled participants, Federal prison, Texas, July 12—August 9, 2021. The timelines of specimen collection and laboratory results for 95 included participants are represented diagrammatically, indexed by the day of onset. Onset was determined to be either a) date of first onset of self-reported symptom(s) meeting the case definition of COVID-19 or b) date of first positive diagnostic SARS-CoV-2 test, whichever occurred first. Each participant is represented by a horizontal line corresponding to the investigation sampling period during their time-course of illness. Participants who were not fully vaccinated (including 2 participants who received only the first dose of a two-dose COVID-19 vaccine series) are depicted at the top of the figure, while fully vaccinated participants are depicted at the bottom. RT-PCR results are represented by solid circles (positive results) or open circles (negative results). For specimens with positive RT-PCR results for which viral culture was performed, culture results are indicated by overlaid blue boxes (positive culture results) or red boxes (negative culture results). Specimens with positive RT-PCR results with a cycle threshold (Ct) value greater than 35 for which viral culture was not performed are indicated by overlaid orange boxes (indicated a presumptive negative viral culture result). Some participants provided specimens during case-finding testing while in quarantine and may have RT-PCR negative specimens collected prior to onset.

On the tenth day of specimen collection, participants were asked to complete a paper-based questionnaire to report COVID-19-like symptoms during the course of their illness, including date of symptom onset and symptom duration. Information on demographic characteristics, COVID-19 vaccination history, previous positive SARS-CoV-2 diagnostic tests, and underlying medical conditions was extracted from BOP electronic medical records for all participants.

### Laboratory Methods

Specimens were collected using nylon flocked minitip swabs, transferred into universal viral transport media (VTM) (Becton Dickinson, Franklin Lakes, NJ) immediately stored at 2-8°C and frozen at -20°C or colder within 72 hours, and sent to CDC for RT-PCR testing using the CDC Influenza SARS-CoV-2 Multiplex Assay. Remnant aliquots were stored at -70°C or below for viral culture. Due to capacity limitations, viral culture was performed on a subset of collected specimens. Specimens were included for viral culture if they had been collected 0, 3, 5, 7, or 9 days since onset and had an accompanying positive RT-PCR test with cycle threshold (Ct) value less than 35. For verification that this selected Ct cutoff did not exclude specimens containing culturable virus, viral culture was also performed on 25 of 102 specimens with Ct>35. (25/25 of these specimens were culture negative.) For more granular detail across the time-course of infection, viral culture was also performed on a subset of specimens collected on other days (see Supplemental Figures 1-2 for details on specimens included for viral culture).

Specimens selected for culture were used to perform limiting-diluting inoculation of Vero CCL-81 cells expressing TMPRSS2, and cultures showing evidence of cytopathic effect were tested by RT-PCR for the presence of SARS-CoV-2 RNA. Viral recovery was as previously described.^10^ Whole genome sequencing (WGS) was performed for one RT-PCR-positive specimen per participant with Ct less than 30 (per sequencing laboratory standard protocols).

### Statistical Methods

Onset (used as time 0 in longitudinal analyses below) was defined to be either a) date of first onset of self-reported symptom(s) meeting the case definition of COVID-19,^11^ or b) date of first positive diagnostic SARS-CoV-2 test, whichever occurred first. In two instances where a participant without symptoms had an initial positive test followed by at least 3 negative tests before subsequent positive tests, the date of second positive test was used.

Participants were considered fully vaccinated if ≥14 days had elapsed since they had completed all recommended doses of a COVID-19 primary vaccine series before the start of the outbreak. (No participant had completed a primary vaccine series <14 days before the outbreak.) Participants were considered not fully vaccinated if they had not received any doses of a vaccine or if they had not completed all doses of a vaccine series. Demographic characteristics of participants stratified by vaccination status were assessed using Fisher’s exact tests.

Three surrogate markers for assessing transmission potential were analyzed as primary outcomes: RT-PCR positivity (an indicator of current/recent infection), RT-PCR Ct value (a semi-quantitative indicator of relative level of viral nucleic acid), and viral culture positivity (an indicator of viable/infectious virus). Dichotomous laboratory results (RT-PCR positivity and viral culture positivity) were analyzed longitudinally with time 0 defined as the date of onset and the primary endpoints defined by a participant’s last positive test. Specimens for which viral culture was not performed were presumed to be culture negative if an accompanying RT-PCR test was negative or was positive with Ct>35. To account for variation in the interval between onset and enrollment, and intermittent participation in specimen collection by some participants (which can result in interval and right censoring), survival analyses were performed using Turnbull estimation using the “interval” package implementation in R.^12^ Hypothesis testing of survival functions was performed using the generalized Wilcoxon-Mann-Whitney method for interval-censored data.

As a post-hoc evaluation of potential interactions between vaccination status and known prior SARS-CoV-2 infections, a stratified analysis was conducted using Fisher’s exact test to compare RT-PCR and viral culture results across these two variables among specimens collected on days with complete viral culture coverage (0, 3, 5, 7, and 9 days since onset).

Non-dichotomous laboratory results (RT-PCR Ct values) were characterized by days since onset using medians and interquartile ranges (IQRs). Because Ct values are semi-parametric, distributions were compared non-parametrically using the Mann-Whitney U test with ties (for dichotomous variables) or the Kruskal-Wallis test (for categorical variables with more than 2 levels); negative RT-PCR results were assigned higher ranks than any Ct value from positive RT-PCR results. To account for multiple hypothesis testing across days, α thresholds were adjusted using Bonferroni correction. All hypothesis tests performed are detailed in Supplementary Tables 1 and 2. All statistical analyses were performed in R version 4.0.2 (R Core Team, Vienna, Austria).

## Results

### Population Characteristics

Among 189 persons with SARS-CoV-2 infection eligible to enroll, a total of 96 persons consented to participate in serial specimen collection; one participant had a single positive diagnostic test (Ct=36.2) followed by seven negative diagnostic tests and reported no symptoms and was excluded as a non-case. Of the 95 included participants, 78 (82%) were documented as being fully vaccinated against SARS-CoV-2, 15 (16%) were unvaccinated and 2 (2%) were partially vaccinated and categorized as not fully vaccinated in further analyses (Table 1). Among fully vaccinated participants, a majority (57/78, 73%) received the Pfizer vaccine; smaller proportions received the Moderna vaccine (14/78, 18%) or Janssen vaccine (7/78, 9%). A majority (47/78, 60%) of fully vaccinated participants completed their vaccination series more than 120 days prior to the start of the outbreak (IQR: 81-140 days prior to start). Recipients of Pfizer vaccines completed their series earlier (IQR: 131-131 days) than recipients of Moderna (IQR: 81-82 days prior to start) or Janssen (IQR: 46-70 days prior to start) vaccines (p<0.001). A small number of participants (2/78 fully vaccinated, 3%, and 2/17 not fully vaccinated, 12%, p=0.10) had a documented prior SARS-CoV-2 infection. Based on symptom self-report at the end of sampling, 76% of participants reported at least one symptom in the COVID-19 case definition [CSTE 2021]. The most commonly reported symptoms were runny or stuffy nose (58%), loss of smell or taste (54%), and cough (45%). Of 95 specimens from 95 participants for which sequencing was attempted, 64 were successfully sequenced and passed quality metrics; all 64 (100%) belonged to the B.1.617.2 (Delta) lineage and AY.3 sublineage.

**Table 1.**
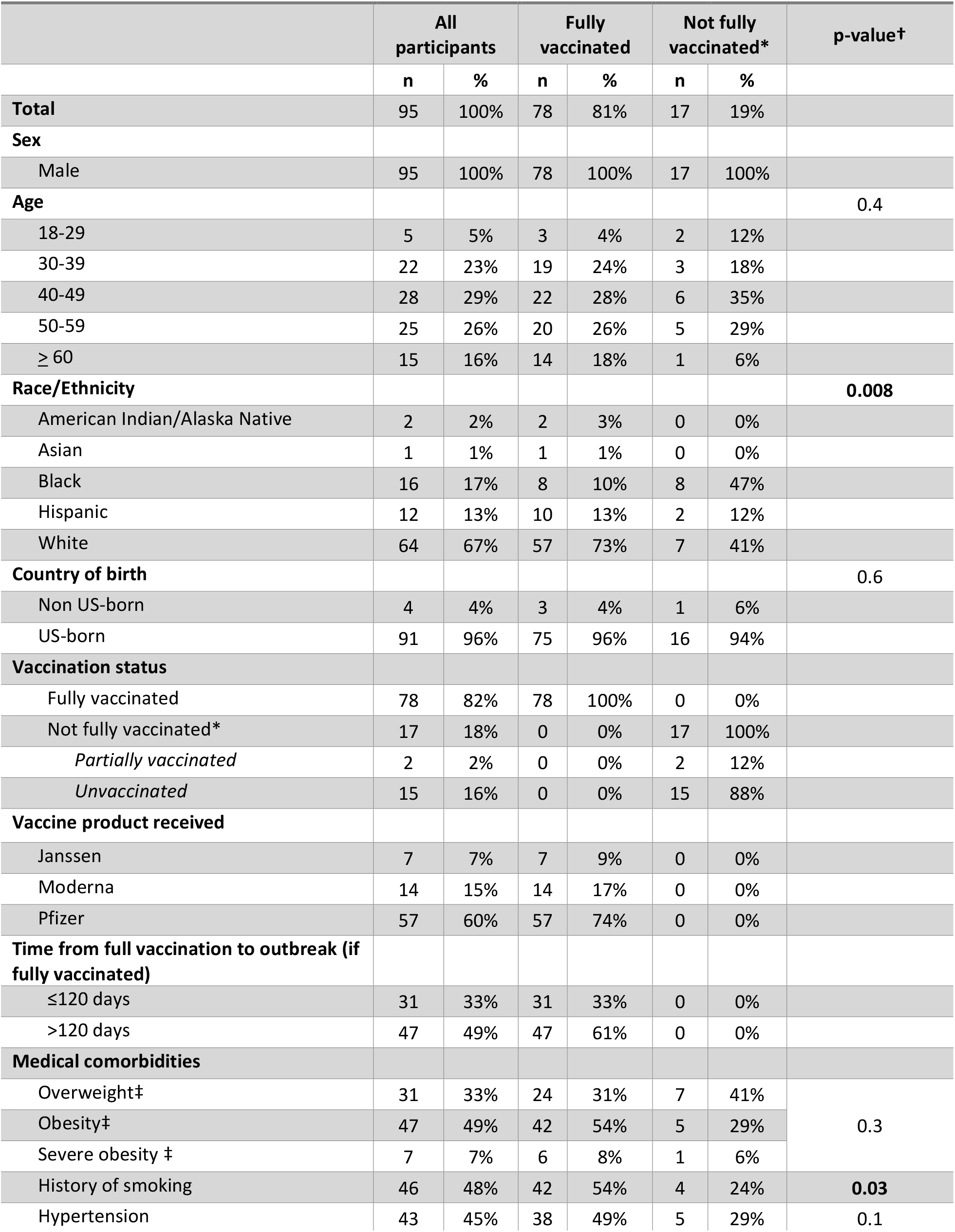
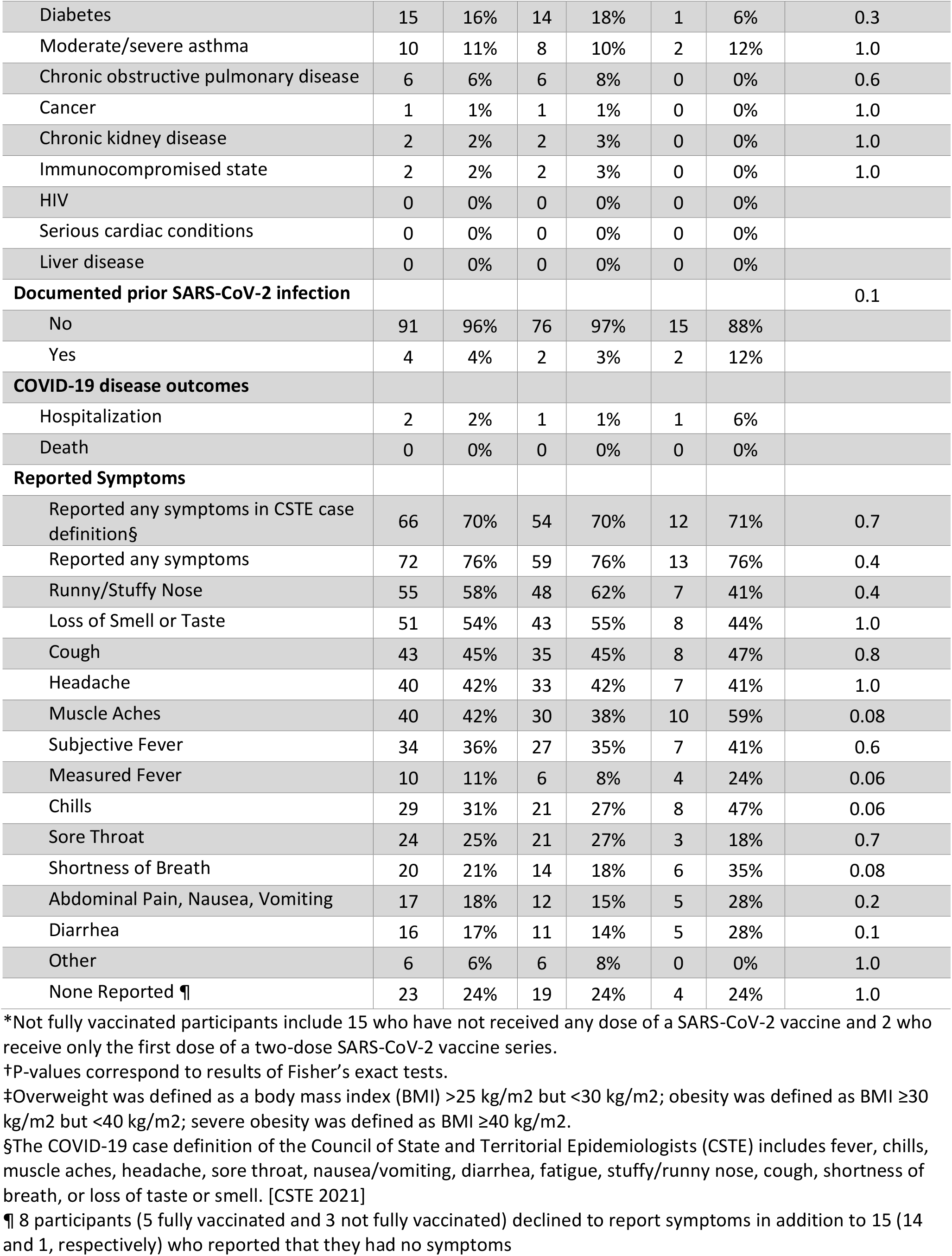
Characteristics of enrolled participants who tested positive for SARS-CoV-2, Federal prison, Texas, July 12—August 9, 2021.

### RT-PCR Positivity

From the 95 included participants, 978 specimens were collected for RT-PCR testing (825/978, 84% from fully vaccinated participants). Specimens were collected ranging from 13 days prior to onset (among participants tested during quarantine prior to diagnosis) to 32 days following onset. See Figure 1 for a diagrammatic representation of RT-PCR specimen collection from participants, and see Supplemental Figure 1 for details of specimen collection by day since onset (stratified by vaccination status). A median of 6 days elapsed between onset and enrollment among fully vaccinated participants, compared with a median of 7 days among participants who were not fully vaccinated (p=0.33). Overall, 499 of the 978 (51%) specimens tested positive by RT-PCR.

No significant differences in time to last RT-PCR positive test were found. Median duration of RT-PCR positivity was 13 days among fully vaccinated participants versus 13 days among participants who were not fully vaccinated (p=0.50; Figure 2); and 10 days among participants with known history of prior SARS-CoV-2 infection (regardless of vaccination) versus 13 days among participants without any known prior infection (p=0.12). Among fully vaccinated participants, median duration of positivity was 10 days among Moderna vaccine recipients versus 13 days among Pfizer recipients and 13 days among Janssen recipients (p=0.39); and 13 days among participants fully vaccinated more than 120 days prior to the outbreak versus 11 days among participants vaccinated 120 days or less prior to the outbreak (p=0.32).

**Figure 2.**
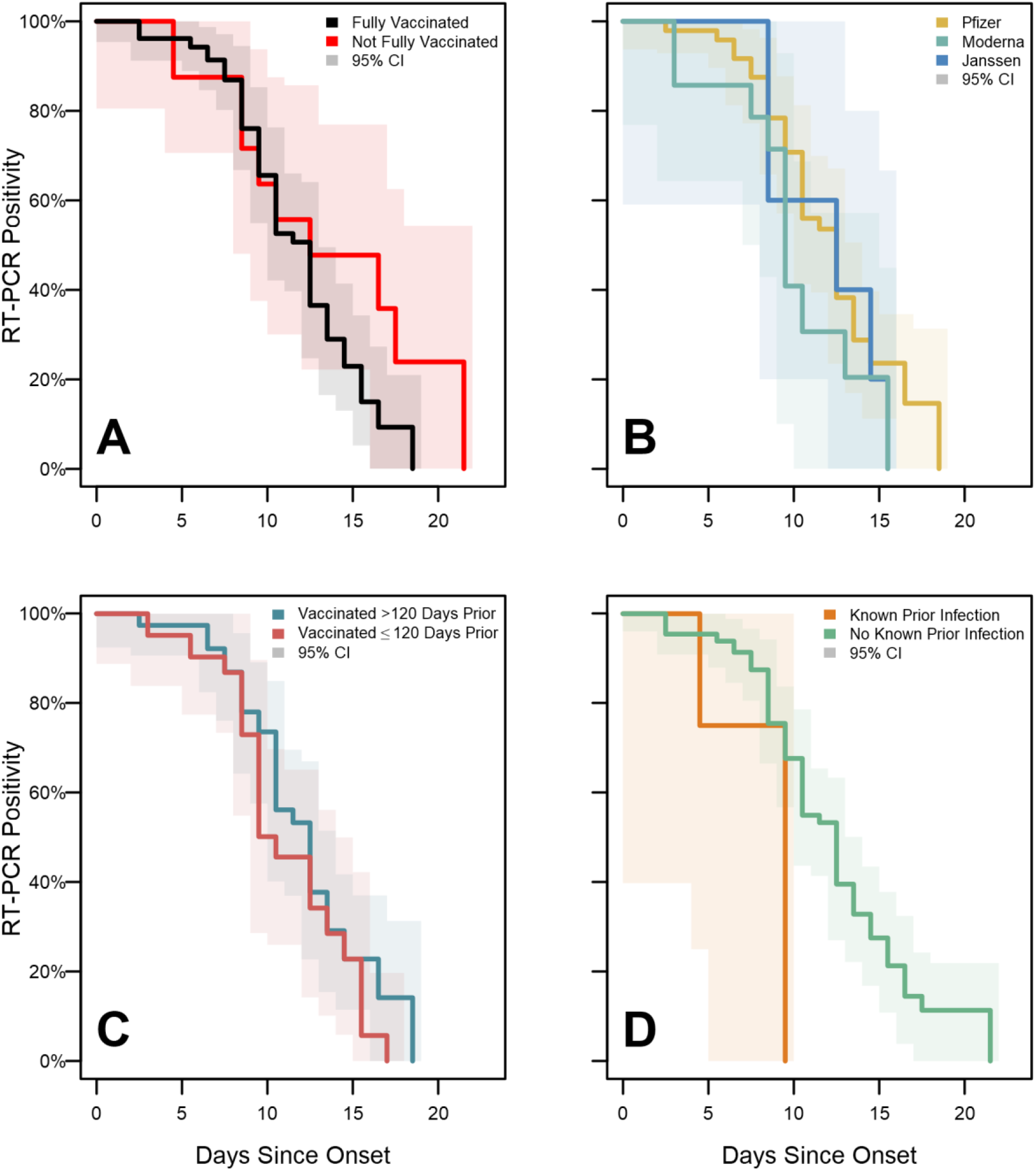
SARS-CoV-2 RT-PCR test positivity survival curves for enrolled participants, Federal prison, Texas, July 12—August 9, 2021. Panels illustrate the results of Turnbull estimation survival functions with a primary endpoint of last positive reverse transcription-polymerase chain reaction (RT-PCR) test result. Solid lines indicate nonparametric maximum likelihood estimates and shaded regions correspond to 95% confidence intervals estimated through modified bootstrap. Survival functions are plotted by Turnbull interval midpoints. Onset was determined to be either a) date of first onset of self-reported symptom(s) meeting the case definition of COVID-19 or b) date of first positive diagnostic SARS-CoV-2 test, whichever occurred first. Panel A depicts RT-PCR positivity by vaccination status (not fully vaccinated participants include 2 participants who received only the first dose of a two-dose COVID-19 vaccine series). Panel B depicts positivity by vaccine product among fully vaccinated participants. Panel C depicts positivity according to the time from completion of a COVID-19 vaccine/series to onset. Panel D depicts positivity according to history of known prior SARS-CoV-2 infection.

### Ct Values

Ct values from specimens testing positive by RT-PCR increased with the number of days since onset (Figure 3). Among specimens from fully vaccinated participants, Ct values increased from a median of 26.4 (IQR: 23.5-28.4) on the day of onset to a median of 32.9 on day 10 (IQR: 30.5-34.6), while Ct values from specimens from participants who were not fully vaccinated increased from a median of 28.5 (IQR:24.8-31.8) on the day of onset to a median of 34.5 on day 10 (IQR: 29.4-35.2). Across the time-course of infection, no statistically significant difference was observed among Ct values by vaccination status on any day after Bonferroni correction (all p>0.0026, the Bonferroni-corrected α threshold). Additionally, no significant differences were observed among Ct values when stratified by vaccine product, time since vaccination, or known prior SARS-CoV-2 infection. While not statistically significant, lower Ct values were observed early in the time-course of infection among Janssen vaccine recipients (day 3 median: 17.9; IQR: 17.6-19.4) than among Moderna (day 3 median: 27.4; IQR: 23.7-28.1) or Pfizer recipients (day 3 median: 24.8; IQR: 23.1-26.8; p=0.016 while Bonferroni α=0.0026).

**Figure 3.**
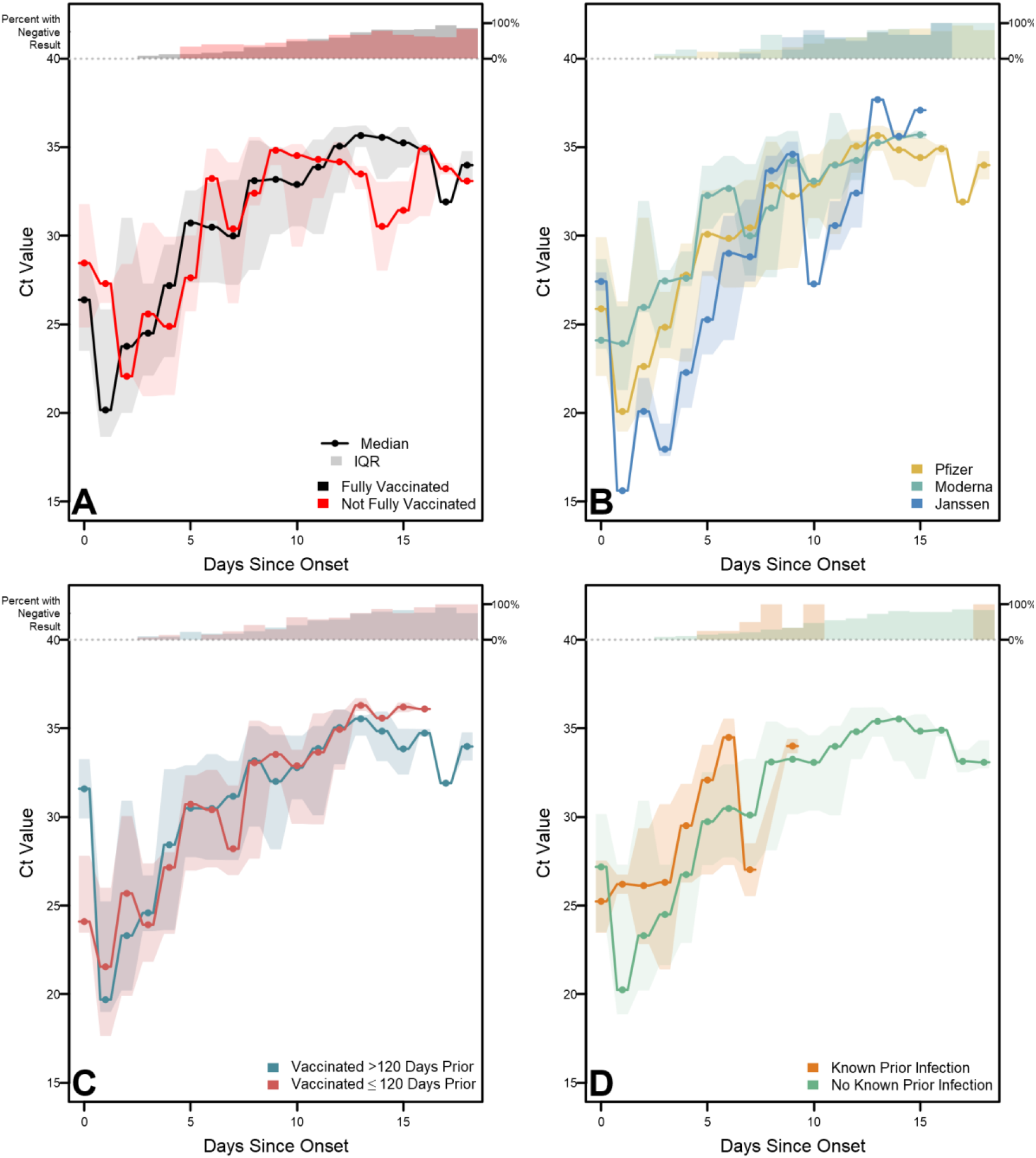
RT-PCR Cycle Threshold distributions for enrolled participants with confirmed SARS-CoV-2 infection, Federal prison, Texas, July 12—August 9, 2021. Panels illustrate daily medians and interquartile ranges (IQRs) for reverse transcription-polymerase chain reaction (RT-PCR) cycle threshold (Ct) values among specimens with positive RT-PCR results. Solid lines indicate median Ct values and shaded regions indicate IQRs. Percentages at the top of each panel indicate the proportion of specimens with negative RT-PCR results each day Onset was determined to be either a) date of first onset of self-reported symptom(s) meeting the case definition of COVID-19 or b) date of first positive diagnostic SARS-CoV-2 test, whichever occurred first. Panel A depicts RT-PCR positivity by vaccination status (not fully vaccinated participants include 2 participants who received only the first dose of a two-dose COVID-19 vaccine series). Panel B depicts positivity by vaccine product among fully vaccinated participants. Panel C depicts positivity according to the time from completion of a COVID-19 vaccine/series to onset. Panel D depicts positivity according to history of known prior SARS-CoV-2 infection.

### Viral Culture Positivity

Of the 978 specimens collected, viral culture was performed on 286 (29%); an additional 556 (57%) were included as presumptive negative viral culture results due to an accompanying negative RT-PCR test (n=479) or a positive RT-PCR test with a Ct value greater than 35 (n=77). Viral culture capture by day since onset stratified by vaccination status is detailed in Supplementary Figure 2. Among the 842 specimens with a viral culture result, 75 (9%) had a positive viral culture. Virus was recovered from 57/690 (8%) of specimens from fully vaccinated participants, compared with 18/152 (12%) of specimens from participants who were not fully vaccinated (p=0.16).

No statistically significant difference was detected in the duration of viral culture positivity (Figure 4) between participants who were fully vaccinated (median: 5 days) compared with those who were not fully vaccinated (median: 5 days; p=0.29). (Viral culture results are illustrated as a function of days since onset and grouped by RT-PCR result in Supplementary Figure 4). Cumulative hazard functions indicate overall shorter culture positivity for fully vaccinated participants who received the Moderna vaccine than those who received Pfizer (p=0.048) or Janssen vaccines (p=0.003), but there was no significant difference between recipients of Pfizer and Janssen vaccines (p=0.12). No statistically significant differences in duration of culture positivity were detected when stratified according to time since vaccination (p=0.79) or known prior infection (p=0.99).

**Figure 4.**
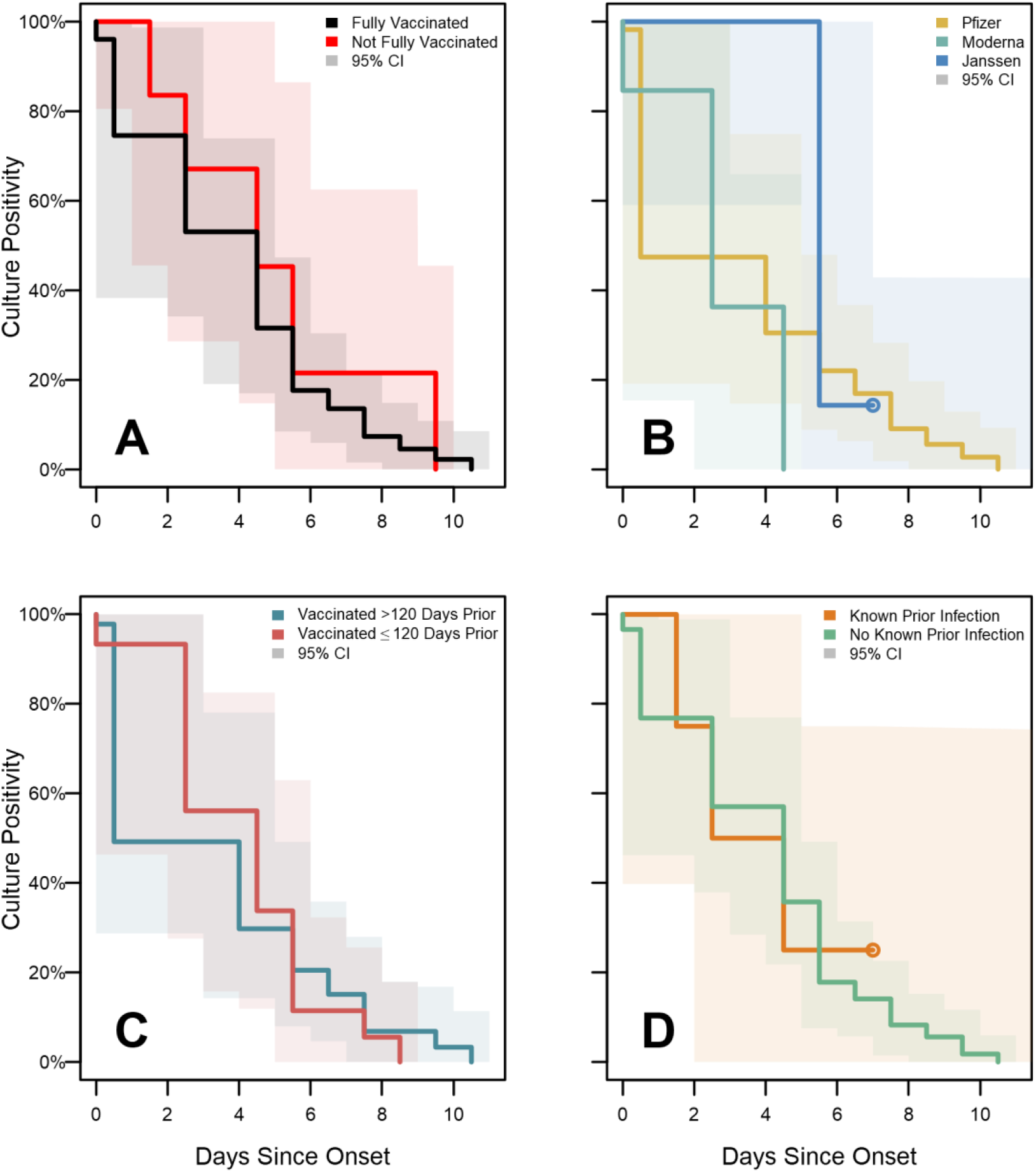
SARS-CoV-2 viral culture test positivity survival curves for enrolled participants, Federal prison, Texas, July 12—August 9, 2021. Panels illustrate the results of Turnbull estimation survival functions with a primary endpoint of last positive viral culture test result. Specimens were included as presumptive negative results if no culture was performed but were accompanied by negative RT-PCR results or positive RT-PCR results with Ct>35. Solid lines indicate nonparametric maximum likelihood estimates and shaded regions correspond to 95% confidence intervals estimated through modified bootstrap. Survival functions are plotted by Turnbull interval midpoints. When Turnbull intervals are bounded by positive infinity (resulting from right-censoring in subgroups), survival functions are truncated by open points at the rightmost non-infinite intervals. Onset was determined to be either a) date of first onset of self-reported symptom(s) meeting the case definition of COVID-19 or b) date of first positive diagnostic SARS-CoV-2 test, whichever occurred first. Panel A depicts RT-PCR positivity by vaccination status (not fully vaccinated participants include 2 participants who received only the first dose of a two-dose COVID-19 vaccine series). Panel B depicts positivity by vaccine product among fully vaccinated participants. Panel C depicts positivity according to the time from completion of a COVID-19 vaccine/series to onset. Panel D depicts positivity according to history of known prior SARS-CoV-2 infection.

### Factorial Stratification: Vaccination Status and History of Prior Infection

Figure 5 illustrates a post-hoc stratification of RT-PCR and viral culture results by vaccination status and prior SARS-CoV-2 infection. No statistically significant difference in RT-PCR or viral culture positivity was detected on any day; however, bivariate stratification resulted in small population sizes in some groups (n=2 participants each for those fully vaccinated with a known prior infection and those not fully vaccinated with a known prior infection), which limits the ability to draw conclusions about these groups.

**Figure 5.**
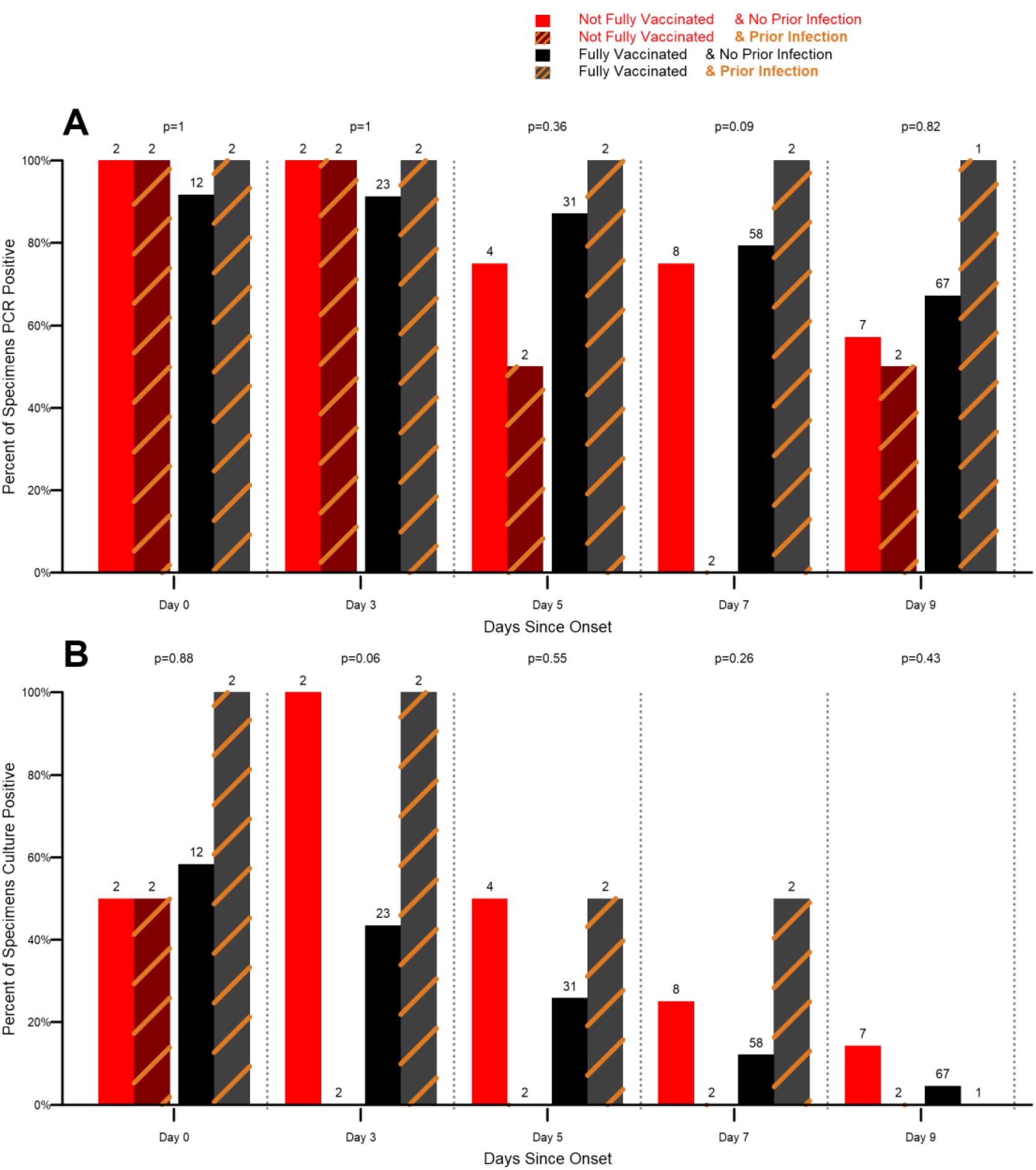
SARS-CoV-2 RT-PCR test positivity (A) and viral culture test positivity (B) stratified by vaccination status and prior infection status for enrolled participants, Federal prison, Texas, July 12— August 9, 2021. Panels illustrate the proportions of specimens for which RT-PCR test results (panel A) or viral culture test results (panel B) were positive, stratified by both vaccination status and history of prior SARS-CoV-2 infection. Solid bars indicate results for participants with no known prior infections, and striped bars indicate results for participants with documented prior infections. Specimens were included as presumptive negative results if no culture was performed but were accompanied by negative RT-PCR results or positive RT-PCR results with Ct>35. Onset was determined to be either a) date of first onset of self-reported symptom(s) meeting the case definition of COVID-19 or b) date of first positive diagnostic SARS-CoV-2 test, whichever occurred first. Results are depicted only for days 0, 3, 5, 7, and 9 since onset, representing days for which 100% of eligible specimens had viral culture performed. Bar labels indicate the number of specimens collected from participants in each group for each day. P-values are reported at the top of each daily grouping and correspond to Fisher’s exact test of proportions across the four groups.

## Discussion

During a high-transmission outbreak of the SARS-CoV-2 Delta variant in a prison setting, we failed to find different durations of RT-PCR positivity, Ct values, or durations of viral culture positivity in fully vaccinated persons compared with persons who were not fully vaccinated. However, vaccinated persons who received the Moderna vaccine had a shorter duration of culture positivity compared with Pfizer or Janssen vaccine recipients. (However, Moderna vaccine recipients also were more recently vaccinated than Pfizer vaccine recipients.) Collectively, our findings suggest that, as evidence continues to emerge in this developing field, vaccinated persons who become infected should be regarded as not significantly less infectious than unvaccinated persons for the purposes of public health action.

As viral infections in vaccinated persons can result from either a failure to mount a protective immune response following initial vaccination or a gradual waning of immunological protection following initially robust protection, the infectiousness of vaccinated persons may be variable. It is plausible that some participants in this investigation who became infected despite vaccination had weak or waning vaccine-induced protection and were therefore similar to unvaccinated persons in the markers of transmission potential that we evaluated.

This report adds to a limited body of scientific literature evaluating the transmission potential of SARS-CoV-2 infections in vaccinated persons. Reports of infections in vaccinated persons have found mixed results using markers of transmission potential, and no longitudinal studies of viral culture characteristics in vaccinated persons with Delta infections have been published. A multi-site serial testing investigation involving Alpha (B.1.1.7) and Gamma (P.1) infections found that duration of culture positivity was shorter among vaccinated persons compared with unvaccinated persons.^13, 14^ One report using surveillance data found lower Ct values among unvaccinated persons, but this difference was only observed for two of three RT-PCR probes and only during one of three months.^15^ One cross-sectional report found no difference in Ct value by vaccination status.^2^ However, extrapolating from cross-sectional and surveillance data may be challenging without data to account for timing of specimen collection in the course of infection. Nevertheless, this finding is corroborated by analysis of a clinical convenience sample which found vaccination did not impact Ct values and reduced viral recovery of Alpha variant but did not reduce recovery of Delta variant virus;^16^ similar findings were mirrored by two retrospective health-system cohorts.^17, 18^ A report of health system workers found that viral culture positivity was reduced in vaccinated persons despite similar Ct values as those in unvaccinated persons.^3^ A separate report found that early in the clinical course of infection, Ct values were comparable between vaccinated and unvaccinated persons, but among individuals who presented to care later in their course of illness, Ct values were higher in vaccinated persons.^19^ A study of household transmission of Delta infections found similar peak viral loads regardless of vaccination status, but noted faster declines in vaccinated persons.^20^ Cumulatively, available data have not clearly or consistently identified markers of reduced transmission potential in vaccinated persons with SARS-CoV-2 infection. This report, which to our awareness represents the first longitudinal investigation of viral culture characteristics of vaccinated persons with Delta variant infections, further demonstrates the potential of vaccinated persons to contribute to SARS-CoV-2 transmission.

While our investigation did not find evidence of reduced transmission potential from vaccinated persons with infection, vaccination is known to reduce the risk of infection,^6, 21^ which prevents secondary transmission. In addition, vaccination remains a strongly protective factor against morbidity and mortality due to SARS-CoV-2.^22^ Protection against infection, morbidity, and mortality underscores the importance of maximizing vaccination coverage, particularly in settings where challenges to physical distancing can result in rapid, widespread transmission when infections do occur.

The evidence that vaccinated persons can transmit SARS-CoV-2 to others suggests that there is continued risk of widespread outbreaks when the virus is introduced into congregate settings, even when vaccination coverage is high. In particular, because of the potential for rapid transmission and high prevalence of underlying health conditions in incarcerated populations,^7, 8^ persons living or working in correctional facilities should quarantine after exposure to SARS-CoV-2, regardless of vaccination status. Post-exposure quarantine is especially important where the risk of transmission is high (e.g., in dorm-style housing, and where staff and/or incarcerated persons frequently interact across housing units) or where the population is at high risk of severe outcomes from COVID-19. Facilities can continue to minimize the need for quarantine by enforcing consistent indoor masking to the extent possible, continuing recommended disinfection, cleaning, and ventilation, and maintaining routine test-based screening programs that can identify cases early and facilitate timely action (including isolation) to limit exposure to others. Facilities that implement routine test-based screening should continue to include vaccinated persons in their frame.

This report is subject to several limitations. Due to the small proportion of participants who were not fully vaccinated (19%), statistical comparisons on the basis of vaccination status were underpowered, and negative findings reported here warrant cautious interpretation. To increase the sample size of this group, two partially vaccinated participants were included, potentially diluting the characteristics of unvaccinated participants. However, our conclusions did not change when analyses were performed excluding these two participants. Similarly, only four participants had known prior infection, of which a higher proportion occurred in those not fully vaccinated; therefore, these participants may appear to have slightly greater immunological protection than those without prior infection. On average, unvaccinated participants enrolled earlier in the outbreak and later in their course of infection than vaccinated participants; we utilized Turnbull estimation in survival analyses to account for the possibility of interval censoring in this population. All symptom data was self-reported and collected at the end of the specimen collection period, which may have impacted the accuracy of participants’ recall related to the date of symptom onset. Ct values are semi-quantitative indicators of viral RNA levels and cannot be interpreted as quantitative markers of viral load or infectiousness. To avoid drawing quantitative conclusions around Ct values, we conservatively utilized non-parametric rank-based statistics (Mann-Whitney and Kruskal-Wallis) with Bonferroni correction to describe Ct values in this investigation. Information on prior SARS-CoV-2 infection was obtained from medical records; persons with earlier infections that were undiagnosed or diagnosed prior to incarceration and not documented in the BOP medical system may not have been correctly characterized. Finally, we did not attempt viral culture for 561 specimens with Ct>35 and classified them as presumptively negative. This decision was based on negative viral culture results from 25/25 specimens with Ct>35 for which viral culture was performed during this investigation, as well as previously published findings demonstrating an inability to recover viable virus from specimens that were RT-PCR negative.^23^

In this investigation, we found no statistically significant difference in transmission potential between vaccinated persons and persons who were not fully vaccinated. Therefore, our findings indicate that prevention and mitigation measures should be applied without regard to vaccination status for persons in high-risk settings or those with significant exposures. In congregate settings, and correctional and detention facilities in particular, post-exposure testing and quarantine remain essential tools to limit transmission when cases are identified, in addition to other recommended prevention measures.^24^ Our data add to a growing body of evidence characterizing transmission potential from vaccinated persons. Future studies of transmission potential from vaccinated persons with infection, incorporating similar laboratory-based markers as well as evidence of transmission from secondary attack rates and network analysis, may help to further describe the contributions of vaccinated persons in chains of transmission as the pandemic evolves and new variants emerge.

## Data Availability

Deidentified data used in the present analysis may be made available upon reasonable request to the authors

## Acknowledgements

Mario Cordova, Torrey Haskins, Jennifer Jackson, Joshua Jett, Barbara Swopes, Tammy Winbush, Federal Bureau of Prisons.

## Footnotes

* 45 C.F.R. part 46, 21 C.F.R. part 56; 42 U.S.C. Sect. 241(d); 5 U.S.C. Sect. 552a; 44 U.S.C. Sect. 3501 et seq.

## References

1. Mancuso M, Eikenberry SE, Gumel AB. Will vaccine-derived protective immunity curtail COVID-19 variants in the US? Infect Dis Model. 2021; 6: 1110–34. 10.1016/j.idm.2021.08.008.

2. Riemersma KK, Grogan BE, Kita-Yarbro A, Halfmann PJ, Segaloff HE, Kocharian A, et al. Shedding of Infectious SARS-CoV-2 Despite Vaccination. medRxiv. 2021: 2021.07.31.21261387. 10.1101/2021.07.31.21261387.

3. Shamier MC, Tostmann A, Bogers S, de Wilde J, IJpelaar J, van der Kleij WA, et al. Virological characteristics of SARS-CoV-2 vaccine breakthrough infections in health care workers. medRxiv. 2021: 2021.08.20.21262158. 10.1101/2021.08.20.21262158.

4. Brown CM, Vostok J, Johnson H, Burns M, Gharpure R, Sami S, et al. Outbreak of SARS-CoV-2 Infections, Including COVID-19 Vaccine Breakthrough Infections, Associated with Large Public Gatherings -Barnstable County, Massachusetts, July 2021. MMWR Morb Mortal Wkly Rep. 2021; 70(31): 1059–62. 10.15585/mmwr.mm7031e2.

5. Centers for Disease Control and Prevention. COVID Data Tracker. Atlanta, GA. US Department of Health and Human Services. Accessed October 16, 2021. [Available from: https://covid.cdc.gov/covid-data-tracker/#trends_dailycases.

6. Hagan LM, McCormick DW, Lee C, Sleweon S, Nicolae L, Dixon T, et al. Outbreak of SARS-CoV-2 B.1.617.2 (Delta) Variant Infections Among Incarcerated Persons in a Federal Prison - Texas, July-August 2021. MMWR Morb Mortal Wkly Rep. 2021; 70(38): 1349–54. 10.15585/mmwr.mm7038e3.

7. Hagan LM, Williams SP, Spaulding AC, Toblin RL, Figlenski J, Ocampo J, et al. Mass Testing for SARS-CoV-2 in 16 Prisons and Jails - Six Jurisdictions, United States, April-May 2020. MMWR Morb Mortal Wkly Rep. 2020; 69(33): 1139–43. 10.15585/mmwr.mm6933a3.

8. Maruschak L, Bronson J, Alper M. Medical problems reported by prisoners, survey of prison inmates, 2016. Washington, DC. US Department of Justice, Bureau of Justice Statistics. 2021. Available from: https://bjs.ojp.gov/sites/g/files/xyckuh236/files/media/document/mprpspi16st.pdf.

9. McMichael TM, Clark S, Pogosjans S, Kay M, Lewis J, Baer A, et al. COVID-19 in a Long-Term Care Facility - King County, Washington, February 27-March 9, 2020. MMWR Morb Mortal Wkly Rep. 2020; 69(12): 339–42. 10.15585/mmwr.mm6912e1.

10. Harcourt J, Tamin A, Lu X, Kamili S, Sakthivel SK, Murray J, et al. Severe Acute Respiratory Syndrome Coronavirus 2 from Patient with Coronavirus Disease, United States. Emerg Infect Dis. 2020; 26(6): 1266–73. 10.3201/eid2606.200516.

11. Council of State and Territorial Epidemiologists. Update to the standardized surveillance case definition and national notification for 2019 novel coronavirus disease (COVID-19). Accessed October 15, 2021. [Available from: https://cdn.ymaws.com/www.cste.org/resource/resmgr/21-ID-01_COVID-19_updated_Au.pdf.

12. Fay MP, Shaw PA. Exact and Asymptotic Weighted Logrank Tests for Interval Censored Data: The interval R package. J Stat Softw. 2010; 36(2). 10.18637/jss.v036.i02.

13. Ke R, Martinez PP, Smith RL, Gibson LL, Achenbach CJ, McFall S, et al. Longitudinal analysis of SARS-CoV-2 vaccine breakthrough infections reveal limited infectious virus shedding and restricted tissue distribution. medRxiv. 2021. 10.1101/2021.08.30.21262701.

14. Ke R, Martinez PP, Smith RL, Gibson LL, Mirza A, Conte M, et al. Daily sampling of early SARS-CoV-2 infection reveals substantial heterogeneity in infectiousness. medRxiv. 2021. 10.1101/2021.07.12.21260208.

15. Griffin JB, Haddix M, Danza P, Fisher R, Koo TH, Traub E, et al. SARS-CoV-2 Infections and Hospitalizations Among Persons Aged ≥16 Years, by Vaccination Status - Los Angeles County, California, May 1-July 25, 2021. MMWR Morb Mortal Wkly Rep. 2021; 70(34): 1170–6. 10.15585/mmwr.mm7034e5.

16. Luo CH, Morris CP, Sachithanandham J, Amadi A, Gaston D, Li M, et al. Infection with the SARS-CoV-2 Delta Variant is Associated with Higher Infectious Virus Loads Compared to the Alpha Variant in both Unvaccinated and Vaccinated Individuals. medRxiv. 2021. 10.1101/2021.08.15.21262077.

17. Christensen PA, Olsen RJ, Long SW, Subedi S, Davis JJ, Hodjat P, et al. Delta variants of SARS-CoV-2 cause significantly increased vaccine breakthrough COVID-19 cases in Houston, Texas. medRxiv. 2021: 2021.07.19.21260808. 10.1101/2021.07.19.21260808.

18. Eyre DW, Taylor D, Purver M, Chapman D, Fowler T, Pouwels KB, et al. The impact of SARS-CoV-2 vaccination on Alpha & Delta variant transmission. medRxiv. 2021: 2021.09.28.21264260. 10.1101/2021.09.28.21264260.

19. Chia PY, Xiang Ong SW, Chiew CJ, Ang LW, Chavatte J-M, Mak T-M, et al. Virological and serological kinetics of SARS-CoV-2 Delta variant vaccine-breakthrough infections: a multi-center cohort study. medRxiv. 2021: 2021.07.28.21261295. 10.1101/2021.07.28.21261295.

20. Singanayagam A, Hakki S, Dunning J, Madon KJ, Crone M, Koycheva A, et al. Community transmission and viral load kinetics of SARS-CoV-2 Delta (B.1.617.2) variant in vaccinated and unvaccinated individuals. Preprint Available at SSRN: https://ssrncom/abstract=3918287 or http://dxdoiorg/102139/ssrn3918287. 2021.

21. Pouwels KB, Pritchard E, Matthews PC, Stoesser N, Eyre DW, Vihta KD, et al. Effect of Delta variant on viral burden and vaccine effectiveness against new SARS-CoV-2 infections in the UK. Nat Med. 2021. 10.1038/s41591-021-01548-7.

22. Tenforde MW, Patel MM, Ginde AA, Douin DJ, Talbot HK, Casey JD, et al. Effectiveness of SARS-CoV-2 mRNA Vaccines for Preventing Covid-19 Hospitalizations in the United States. Clin Infect Dis. 2021. 10.1093/cid/ciab687.

23. Ford L, Lee C, Pray IW, Cole D, Bigouette JP, Abedi GR, et al. Epidemiologic Characteristics Associated With Severe Acute Respiratory Syndrome Coronavirus 2 (SARS-CoV-2) Antigen-Based Test Results, Real-Time Reverse Transcription Polymerase Chain Reaction (rRT-PCR) Cycle Threshold Values, Subgenomic RNA, and Viral Culture Results From University Testing. Clin Infect Dis. 2021; 73(6): e1348–e55. 10.1093/cid/ciab303.

24. Centers for Disease Control and Prevention. Interim guidance on management of coronavirus disease 2019 (COVID-19) in correctional and detention facilities. Atlanta, GA. US Department of Health and Human Services. Accessed October 29, 2021. [Available from: https://www.cdc.gov/coronavirus/2019-ncov/community/correction-detention/guidance-correctional-detention.html.

