## Supplementary Materials for "Transmission potential of vaccinated and unvaccinated persons infected with the SARS-CoV-2 Delta variant in a federal prison, July—August 2021"

**Supplementary Table 1. Results of hypothesis tests for RT-PCR test viral culture test survival functions.**

| Outcome | Category | Reference Group | Test Statistic* | P-Value* |
|---|---|---|---|---|
| RT-PCR Survival Function | Unvaccinated | Fully Vaccinated | -1.30 | 0.50 |
| RT-PCR Survival Function | Pfizer | Moderna | -2.70 | 0.13 |
| RT-PCR Survival Function | Pfizer | Janssen | -0.06 | 0.97 |
| RT-PCR Survival Function | Moderna | Janssen | 0.65 | 0.62 |
| RT-PCR Survival Function | Pfizer | All Products | -2.48 | 0.39 |
| RT-PCR Survival Function | Moderna | All Products | 2.63 | 0.39 |
| RT-PCR Survival Function | Janssen | All Products | -0.15 | 0.39 |
| RT-PCR Survival Function | More than 120 Days | Less than 120 Days | -2.27 | 0.32 |
| RT-PCR Survival Function | Prior Infection | No Prior Infection | 1.62 | 0.12 |
| Viral Culture Survival Function | Unvaccinated | Fully Vaccinated | -1.91 | 0.29 |
| Viral Culture Survival Function | Pfizer | Moderna | -2.84 | 0.048 |
| Viral Culture Survival Function | Pfizer | Janssen | 1.60 | 0.12 |
| Viral Culture Survival Function | Moderna | Janssen | 3.01 | 0.003 |
| Viral Culture Survival Function | More than 120 Days | Less than 120 Days | -0.64 | 0.79 |
| Viral Culture Survival Function | Prior Infection | No Prior Infection | 0.00 | 0.99 |

*****Test statistics and p-values correspond to the generalized Wilcoxon-Mann-Whitney hypothesis testing method for interval-censored data.

**Supplementary Table 2 . Results of hypothesis tests for Cycle threshold value distributions.**

| Category | Days Since Onset | Test Name | Test Statistic | Bonferroni α | P-Value |
|---|---|---|---|---|---|
| **Vaccination Status** | 0 | Wilcoxon Mann-Whitney U | 0.21 | 0.0026 | 0.832 |
| **Vaccination Status** | 1 | Wilcoxon Mann-Whitney U | 1.34 | 0.0026 | 0.180 |
| **Vaccination Status** | 2 | Wilcoxon Mann-Whitney U | 0.35 | 0.0026 | 0.729 |
| **Vaccination Status** | 3 | Wilcoxon Mann-Whitney U | 0.13 | 0.0026 | 0.899 |
| **Vaccination Status** | 4 | Wilcoxon Mann-Whitney U | -0.82 | 0.0026 | 0.412 |
| **Vaccination Status** | 5 | Wilcoxon Mann-Whitney U | 0.31 | 0.0026 | 0.755 |
| **Vaccination Status** | 6 | Wilcoxon Mann-Whitney U | 1.53 | 0.0026 | 0.127 |
| **Vaccination Status** | 7 | Wilcoxon Mann-Whitney U | 1.10 | 0.0026 | 0.272 |
| **Vaccination Status** | 8 | Wilcoxon Mann-Whitney U | 0.41 | 0.0026 | 0.682 |
| **Vaccination Status** | 9 | Wilcoxon Mann-Whitney U | 1.03 | 0.0026 | 0.301 |
| **Vaccination Status** | 10 | Wilcoxon Mann-Whitney U | 0.54 | 0.0026 | 0.587 |
| **Vaccination Status** | 11 | Wilcoxon Mann-Whitney U | -0.31 | 0.0026 | 0.756 |
| **Vaccination Status** | 12 | Wilcoxon Mann-Whitney U | 0.43 | 0.0026 | 0.665 |
| **Vaccination Status** | 13 | Wilcoxon Mann-Whitney U | -0.89 | 0.0026 | 0.374 |
| **Vaccination Status** | 14 | Wilcoxon Mann-Whitney U | -0.42 | 0.0026 | 0.672 |
| **Vaccination Status** | 15 | Wilcoxon Mann-Whitney U | -1.20 | 0.0026 | 0.231 |
| **Vaccination Status** | 16 | Wilcoxon Mann-Whitney U | -1.28 | 0.0026 | 0.200 |
| **Vaccination Status** | 17 | Wilcoxon Mann-Whitney U | -1.70 | 0.0026 | 0.090 |
| **Vaccination Status** | 18 | Wilcoxon Mann-Whitney U | -0.13 | 0.0026 | 0.894 |
| **Vaccine Product** | 0 | Kruskal-Wallis | 0.35 | 0.0026 | 0.839 |
| **Vaccine Product** | 1 | Kruskal-Wallis | 2.71 | 0.0026 | 0.258 |
| **Vaccine Product** | 2 | Kruskal-Wallis | 3.07 | 0.0026 | 0.215 |
| **Vaccine Product** | 3 | Kruskal-Wallis | 8.19 | 0.0026 | 0.017 |
| **Vaccine Product** | 4 | Kruskal-Wallis | 6.79 | 0.0026 | 0.034 |
| **Vaccine Product** | 5 | Kruskal-Wallis | 3.86 | 0.0026 | 0.145 |
| **Vaccine Product** | 6 | Kruskal-Wallis | 4.70 | 0.0026 | 0.095 |
| **Vaccine Product** | 7 | Kruskal-Wallis | 0.84 | 0.0026 | 0.658 |
| **Vaccine Product** | 8 | Kruskal-Wallis | 6.20 | 0.0026 | 0.045 |
| **Vaccine Product** | 9 | Kruskal-Wallis | 2.89 | 0.0026 | 0.235 |
| **Vaccine Product** | 10 | Kruskal-Wallis | 2.06 | 0.0026 | 0.356 |
| **Vaccine Product** | 11 | Kruskal-Wallis | 0.12 | 0.0026 | 0.943 |
| **Vaccine Product** | 12 | Kruskal-Wallis | 0.24 | 0.0026 | 0.889 |
| **Vaccine Product** | 13 | Kruskal-Wallis | 0.17 | 0.0026 | 0.916 |
| **Vaccine Product** | 14 | Kruskal-Wallis | 0.31 | 0.0026 | 0.855 |
| **Vaccine Product** | 15 | Kruskal-Wallis | 0.92 | 0.0026 | 0.633 |
| **Vaccine Product** | 16 | Kruskal-Wallis | 1.83 | 0.0026 | 0.400 |
| **Vaccine Product** | 17 | Kruskal-Wallis | 0.14 | 0.0026 | 0.705 |

| | | | | | |
|---|---|---|---|---|---|
| **Vaccine Product** | 18 | Kruskal-Wallis | 0.86 | 0.0026 | 0.353 |
| **Time Since Vaccination** | 0 | Wilcoxon Mann-Whitney U | -1.95 | 0.0026 | 0.052 |
| **Time Since Vaccination** | 1 | Wilcoxon Mann-Whitney U | -0.11 | 0.0026 | 0.916 |
| **Time Since Vaccination** | 2 | Wilcoxon Mann-Whitney U | 0.09 | 0.0026 | 0.929 |
| **Time Since Vaccination** | 3 | Wilcoxon Mann-Whitney U | -0.69 | 0.0026 | 0.488 |
| **Time Since Vaccination** | 4 | Wilcoxon Mann-Whitney U | -0.65 | 0.0026 | 0.516 |
| **Time Since Vaccination** | 5 | Wilcoxon Mann-Whitney U | -1.41 | 0.0026 | 0.158 |
| **Time Since Vaccination** | 6 | Wilcoxon Mann-Whitney U | -0.53 | 0.0026 | 0.595 |
| **Time Since Vaccination** | 7 | Wilcoxon Mann-Whitney U | -0.32 | 0.0026 | 0.746 |
| **Time Since Vaccination** | 8 | Wilcoxon Mann-Whitney U | 1.14 | 0.0026 | 0.255 |
| **Time Since Vaccination** | 9 | Wilcoxon Mann-Whitney U | 0.43 | 0.0026 | 0.669 |
| **Time Since Vaccination** | 10 | Wilcoxon Mann-Whitney U | 1.56 | 0.0026 | 0.119 |
| **Time Since Vaccination** | 11 | Wilcoxon Mann-Whitney U | 0.31 | 0.0026 | 0.754 |
| **Time Since Vaccination** | 12 | Wilcoxon Mann-Whitney U | 0.32 | 0.0026 | 0.750 |
| **Time Since Vaccination** | 13 | Wilcoxon Mann-Whitney U | 0.40 | 0.0026 | 0.686 |
| **Time Since Vaccination** | 14 | Wilcoxon Mann-Whitney U | 0.66 | 0.0026 | 0.510 |
| **Time Since Vaccination** | 15 | Wilcoxon Mann-Whitney U | -0.42 | 0.0026 | 0.671 |
| **Time Since Vaccination** | 16 | Wilcoxon Mann-Whitney U | 1.18 | 0.0026 | 0.239 |
| **Time Since Vaccination** | 17 | Wilcoxon Mann-Whitney U | 0.67 | 0.0026 | 0.500 |
| **Time Since Vaccination** | 18 | Wilcoxon Mann-Whitney U | 1.27 | 0.0026 | 0.204 |
| **Prior Infection** | 0 | Wilcoxon Mann-Whitney U | 0.53 | 0.0026 | 0.595 |
| **Prior Infection** | 1 | Wilcoxon Mann-Whitney U | -0.93 | 0.0026 | 0.352 |
| **Prior Infection** | 2 | Wilcoxon Mann-Whitney U | -0.51 | 0.0026 | 0.613 |
| **Prior Infection** | 3 | Wilcoxon Mann-Whitney U | -0.19 | 0.0026 | 0.849 |
| **Prior Infection** | 4 | Wilcoxon Mann-Whitney U | -0.57 | 0.0026 | 0.569 |
| **Prior Infection** | 5 | Wilcoxon Mann-Whitney U | -0.53 | 0.0026 | 0.594 |
| **Prior Infection** | 6 | Wilcoxon Mann-Whitney U | -0.57 | 0.0026 | 0.569 |
| **Prior Infection** | 7 | Wilcoxon Mann-Whitney U | -0.31 | 0.0026 | 0.760 |
| **Prior Infection** | 8 | Wilcoxon Mann-Whitney U | -2.11 | 0.0026 | 0.035 |
| **Prior Infection** | 9 | Wilcoxon Mann-Whitney U | -0.23 | 0.0026 | 0.819 |
| **Prior Infection** | 10 | Wilcoxon Mann-Whitney U | -1.64 | 0.0026 | 0.101 |
| **Prior Infection** | 18 | Wilcoxon Mann-Whitney U | -0.42 | 0.0026 | 0.676 |

**Supplementary Figure 1. Proportion of participants with specimens collected by day since onset among fully vaccinated and not fully vaccinated participants.**

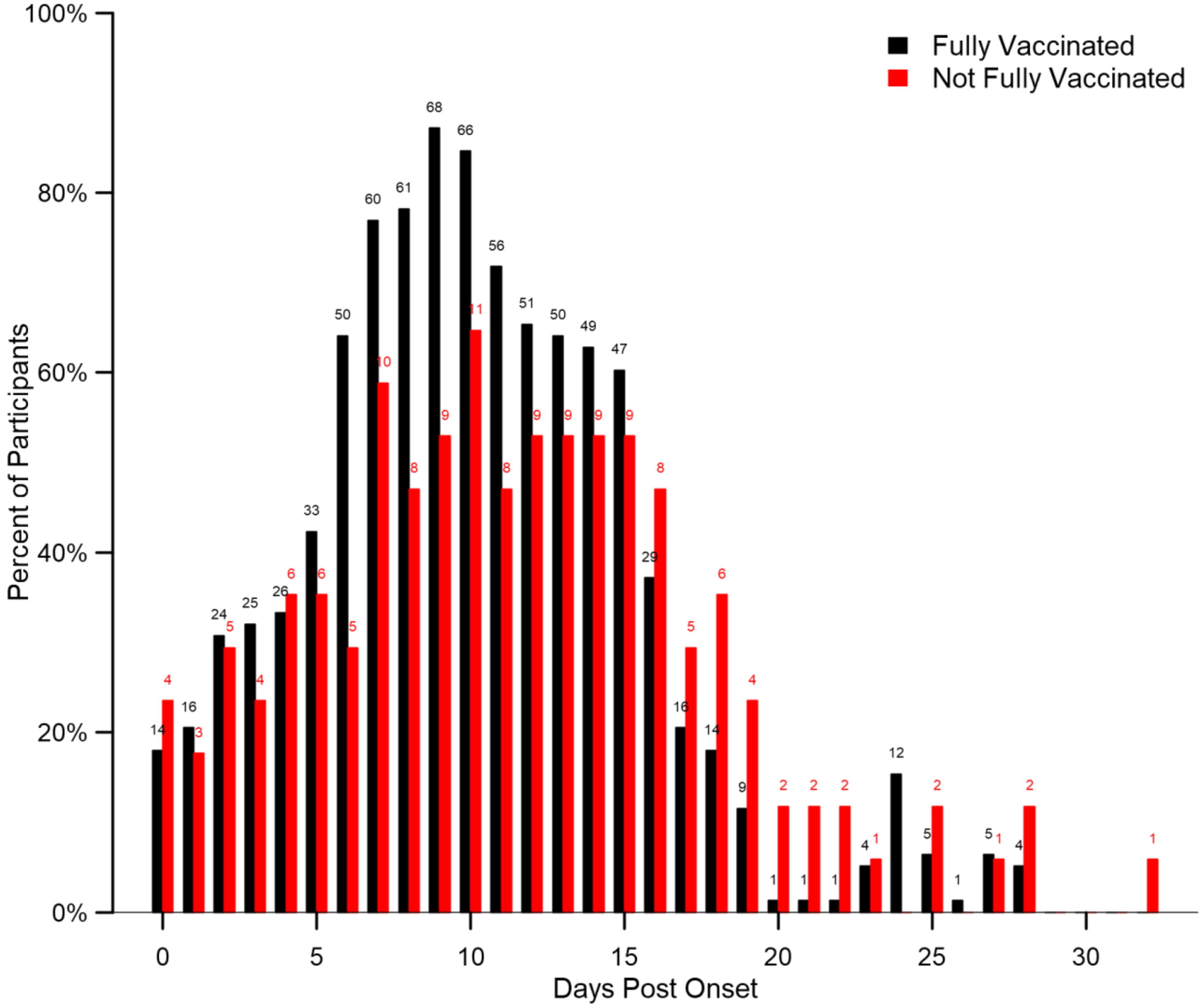


Bar heights indicate the proportion of participants in each group from whom a specimen was collected on a given day since onset. Onset was determined to be either a) date of first onset of self-reported symptom(s) meeting the case definition of COVID-19 or b) date of first positive diagnostic SARS-CoV-2 test, whichever occurred first. Not fully vaccinated participants include 2 participants who received only the first dose of a two-dose COVID-19 vaccine series. Data labels indicate the total number of specimens provided by participants from each group on each day.

**Supplementary Figure 2. Proportion of specimens for which a viral culture result is available among fully vaccinated and not fully vaccinated participants.**

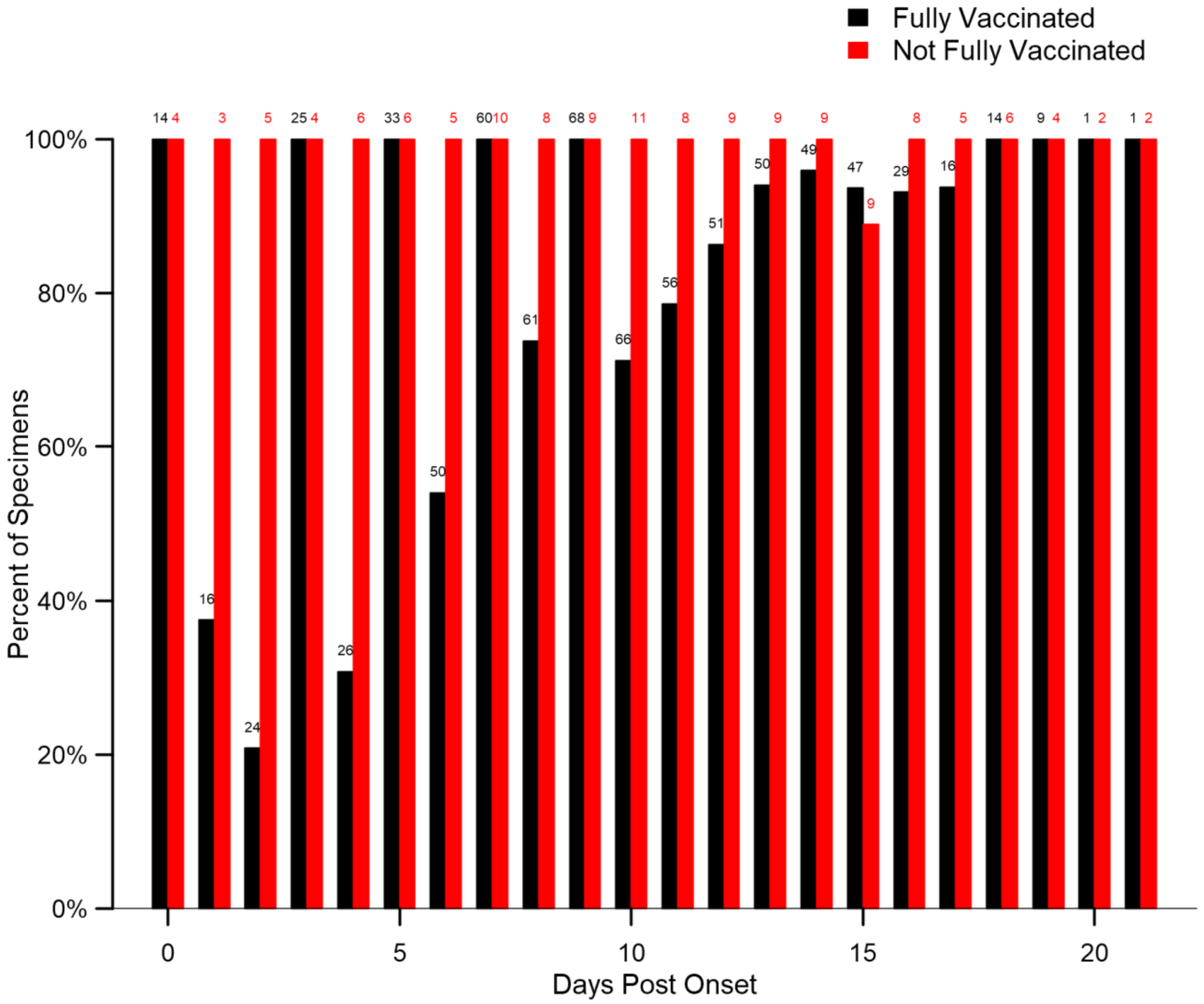


Bar heights indicate the proportion of specimens in each group for which a viral culture result is available on a given day since onset. Specimens for which an accompanying RT-PCR result was negative, or positive with a cycle threshold greater than 35, were presumed to have negative viral culture results and are included here as having a culture result available. Onset was determined to be either a) date of first onset of self-reported symptom(s) meeting the case definition of COVID-19 or b) date of first positive diagnostic SARS-CoV-2 test, whichever occurred first. Not fully vaccinated participants include 2 participants who received only the first dose of a two-dose COVID-19 vaccine series. Data labels indicate the total number of specimens provided by participants from each group on each day.

**Supplementary Figure 3 SARS-CoV-2 Cycle threshold and viral culture test results among fully vaccinated and not fully vaccinated participants among specimens with positive RT-PCR results.**

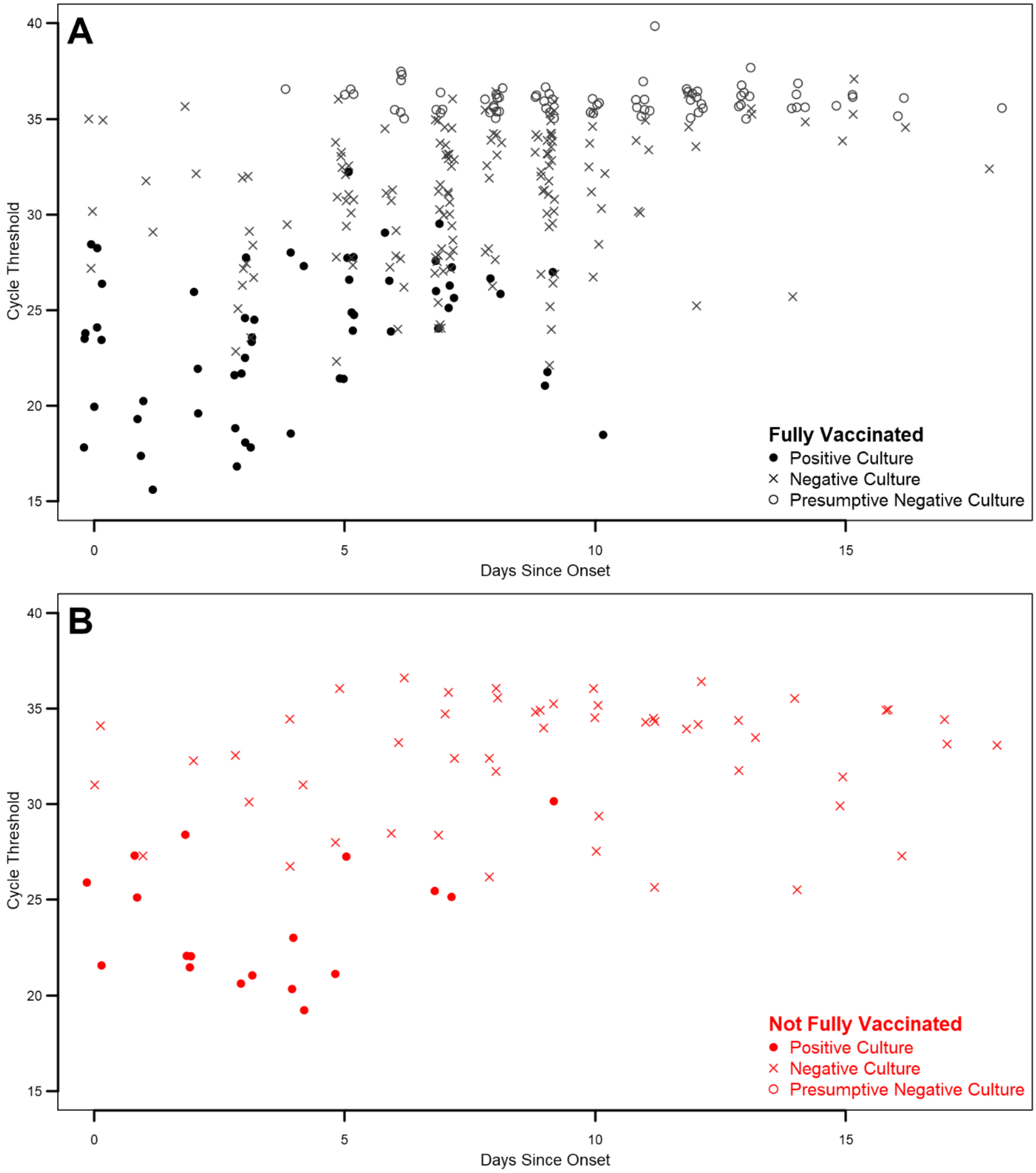


Points illustrate cycle threshold (Ct) values and culture results by specimens collected on a given day since onset. Panel A depicts specimens collected from fully vaccinated participants. Panel B depicts specimens collected from participants who were not fully vaccinated. Specimens with a positive culture result are depicted as closed circles; those with negative culture results are depicted as X's. Specimens with a positive RT-PCR result and Ct value greater than 35 but for which no viral culture was performed were presumed to have a negative viral culture result and are depicted as open circles. Specimens with a negative RT-PCR result and thus no Ct value are not depicted here (but are included in all other analyses as having presumptive negative cultures). X-axis values are jittered to distinguish overlapping points. Onset was determined to be either a) date of first onset of self-reported symptom(s) meeting the case definition of COVID-19 or b) date of first positive diagnostic SARS-CoV-2 test, whichever occurred first. Not fully vaccinated participants include 2 participants who received only the first dose of a two-dose COVID-19 vaccine series.
